# Interactive Evaluation of an Adaptive-Questioning Symptom Checker Using Standardized Clinical Vignettes

**DOI:** 10.1101/2025.08.21.25333628

**Authors:** Prathima Madda, Jagadeesh Kondru

## Abstract

**Objective:** To evaluate the triage performance and history-taking quality of an adaptive-questioning symptom checker (CareRoute) using an interactive protocol on standardized clinical vignettes (Semigran et al., BMJ 2015; 45 cases).

**Methods:** Each session began with only the presenting complaint; CareRoute asked follow-up questions adaptively, and the evaluator answered concisely per the vignette. At the end of questioning, CareRoute issued a triage recommendation. We compared CareRoute’s issued triage with the reference triage and computed history-taking quality from normalized features derived from each vignette’s Condensed Format. History-taking quality comprised (i) elicitation coverage—the percentage of a vignette’s normalized features obtained through questioning, and (ii) elicitation fraction—the proportion of surfaced normalized features (elicited or volunteered) that were obtained through questioning. Primary outcomes were triage concordance and history-taking quality; the secondary outcome was user burden (time spent answering questions). We did not evaluate possible diagnoses, though CareRoute issues them.

**Results:** Exact 3-tier triage concordance was 88.9% (40/45; 95% CI 76.5–95.2%). Elicitation coverage had a median of 67% (IQR 60–71%), and elicitation fraction had a median of 70% (IQR 62–75%). CareRoute asked a median of 19 questions overall (IQR 16–20), with urgency-conditioned questioning: Emergency Care median 10 questions (IQR 4–14), Doctor Visit median 19 questions (IQR 18–20), Self Care median 19 questions (IQR 17–20).

**Conclusions:** In an interactive, vignette-constrained evaluation starting from only the presenting complaint, CareRoute achieved high 3-tier triage concordance (88.9%) with no under-triage on Emergency-reference vignettes, while eliciting most normalized features (median elicitation coverage 67%; median elicitation fraction 70%) with acceptable user burden via urgency-conditioned questioning.

## 1 Introduction

Symptom checkers are consumer-facing digital tools that collect brief medical histories and return preliminary triage advice (and often possible diagnoses). Their practical value hinges on two capabilities: (1) asking the right follow-up questions to surface key findings, and (2) translating those findings into safe, actionable triage guidance. Modern AI-based, adaptive-questioning symptom checkers—such as CareRoute (evaluated in this study)—are designed to elicit missing information and issue a triage recommendation. Evaluations should therefore begin from a presenting complaint and measure both elicitation quality and triage accuracy.

However, most symptom checker evaluations to date use pre-extracted symptom lists entered once, leaving the quality of interactive questioning unmeasured^1–3^. To address this gap and evaluate both questioning quality and triage accuracy in a standardized way, we conducted an interactive evaluation using a well-established set of standardized clinical vignettes. Specifically, we used the 45 standardized clinical vignettes curated by Semigran et al. (BMJ, 2015)^1,4^, which provide validated reference standards and have been widely used in the symptom checker evaluation literature. For brevity, we refer to this dataset as “Semigran-45.”

In Semigran-45, each vignette is labeled with one of three triage categories: *Requires Emergent Care, Requires Non-emergent Care*, or *Self Care appropriate*. Each vignette is also represented in a Condensed Format— a list of findings intended for symptom checker testing^1,4^. We used a normalized version of this list for scoring history-taking quality.

Our study had three objectives: (1) to evaluate CareRoute’s triage accuracy and safety using an interactive protocol that begins with only the presenting complaint, (2) to evaluate CareRoute’s ability to elicit key clinical features through adaptive-questioning, and (3) to establish a reproducible methodology for evaluating history-taking quality in symptom checkers. We pre-specified two primary outcomes: triage concordance (including safety assessment) and history-taking quality (via elicitation coverage and elicitation fraction metrics). We also report user burden (time spent answering questions) as a secondary outcome.

## 2 Methods

We evaluated CareRoute (version 0.3.71) using an interactive, vignette-constrained protocol. Details of the protocol are described below.

### 2.1 Dataset and triage mapping

We used all 45 vignettes from Semigran-45^1,4^ (15 per category: *Requires Emergent Care, Requires Non-emergent Care, Self Care appropriate*). For consistency with CareRoute, we refer to the vignette triage categories as **Emergency Care, Doctor Visit**, and **Self Care**, respectively.

CareRoute provides four triage levels (Emergency Care, Urgent Care, Doctor Visit, Self Care), but our analysis uses a conservative 3-tier mapping that collapses Urgent Care to Doctor Visit.

### 2.2 Condensed Format (official findings list)

For each vignette we started with the Condensed Format list, as published in the Semigran-45 materials, and derived a normalized features list by breaking down composite phrases into discrete features (e.g., “sudden onset severe abdominal pain” into “sudden onset”, “severe”, “abdominal pain”). We treated this normalized list as the set of findings an ideal history should surface for correct triage. The Condensed Format and the normalized features were not shown to CareRoute; they were used only for computing elicitation coverage and elicitation fraction.

#### 2.2.1 Normalized features: example mapping

We normalized composite items from the Semigran-45 Condensed Format into discrete features. For example:

##### Condensed Format of Vignette

> 12 y/o f, sudden onset severe abdominal pain, nausea, vomiting, diarrhea, T=104

##### Normalized features

- 12 y/o f
- sudden onset
- severe
- abdominal pain
- nausea
- vomiting
- diarrhea
- fever 104°F

The normalization process enables granular assessment of history-taking quality via the elicitation coverage and elicitation fraction metrics.

### 2.3 Interactive evaluation protocol

A single physician evaluator (P.M.) completed all 45 vignettes interactively. The evaluator was not presented with the vignette’s reference triage tier, Condensed Format, or normalized features list during questioning. Each session started with only the presenting complaint. CareRoute asked questions adaptively. The evaluator answered to the point, consistent with the vignette. About half of the questions were selection-type; most were multi-select (except yes/no items, which were single-select). Where the vignette included clinician-generated information (e.g., a documented test), the evaluator could report it (e.g., “I was told the test was positive”) when specifically asked by CareRoute. The session ended when CareRoute issued a triage recommendation. Each surfaced normalized feature was labeled *volunteered* if first mentioned before any targeted question about that feature (including in the presenting complaint), and *elicited* if first stated only in response to a targeted question.

### 2.4 Outcomes and statistics

#### Primary outcomes

- Triage concordance with reference values
- Elicitation coverage, 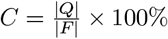
- Elicitation fraction, 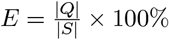

Notation: For each vignette, *F* is the set of normalized features derived from the Condensed Format; *V* are features volunteered by the evaluator (including the presenting complaint); *Q* are features elicited through questioning; *S* = *V* ∪ *Q*.

#### Meaning of the elicitation metrics

*Elicitation coverage* asks: “Of the vignette’s normalized features (the target findings an ideal history should surface), how many did CareRoute obtain by asking questions?” *Elicitation fraction* asks: “Of all surfaced normalized features (volunteered + elicited), what share came from CareRoute’s questions rather than being volunteered?” Higher values on both mean CareRoute’s questioning did more of the work.

#### Secondary outcomes

- User burden (questions asked and estimated time)

#### Reporting standards

Proportions are reported with Wilson 95% confidence intervals^5^. Medians are reported with interquartile ranges (IQRs).

## 3 Results

In the following subsections, we discuss evaluation results in detail. Summarized evaluation results are available in Table A1 (Appendix).

### 3.1 Triage concordance and safety

CareRoute exactly matched the reference in 40/45 cases (88.9%; 95% CI 76.5–95.2%). Under-triage occurred in 1/45 (2.2%) and over-triage in 4/45 (8.9%). On Emergency-reference vignettes, there was no under-triage (15/15; Wilson 95% CI 79.6–100.0%).

#### Characterization of under-triage

The single under-triage (reference: Doctor Visit; CareRoute: Self Care) was an “Influenza” vignette for a 30-year-old woman without risk factors. Current CDC guidance indicates otherwise-healthy adults with uncomplicated influenza can recover at home unless emergency warning signs arise or they belong to higher-risk groups; we therefore regard this as a defensible disagreement with the historical reference rather than a safety concern^6^.

#### Characterization of over-triage

All four over-triage cases were reference *Self Care* vignettes that CareRoute escalated to *Doctor Visit* based on conservative safety cues appropriate for non-urgent outpatient review: infant constipation with intermittent blood-streaked stool and straining; pediatric eczema with sleep disruption and atopy; an upper respiratory illness with ∼6 days of fever and purulent nasal discharge; and acute bronchitis in an older adult with recent fever and a productive cough. These represent safety-favoring shifts to primary care rather than emergency referral.

#### Benchmarking against prior evaluations

Published evaluations report wide variability in triage accuracy for symptom checkers, typically spanning ∼50–90% depending on study design, vignette set, and outcome definitions^2,3^. Within this landscape, CareRoute’s 88.9% exact 3-tier concordance lies near the upper end of reported ranges. Reviews also note that higher-acuity presentations are often triaged more accurately than non-urgent ones, aligning with our finding of no under-triage on Emergency-reference vignettes^2^.

#### 3.1.1 Confusion matrix (rows: Semigran-45 reference; columns: CareRoute results)

| Reference | Emergency Care | Doctor Visit | Self Care | Row total |
| --- | --- | --- | --- | --- |
| Emergency Care (n=15) | **15** | 0 | 0 | 15 |
| Doctor Visit (n=15) | 0 | **14** | 1 | 15 |
| Self Care (n=15) | 0 | 4 | **11** | 15 |
| Column total | 15 | 18 | 12 | 45 |

### 3.2 Elicitation coverage and fraction

Elicitation coverage achieved a median of 67% (IQR 60–71%) across all vignettes, with similar medians across tiers but greater variability for Emergency cases: Emergency 67% (IQR 38–71); Doctor Visit 67% (IQR 65–71); Self Care 67% (IQR 63–73).

Elicitation fraction was 70% (IQR 62–75%). Per-tier medians—Emergency 71.4% (IQR 55.0–75.0), Doctor Visit 66.7% (IQR 64.6–72.1), Self Care 70.0% (IQR 66.7–76.4)—tracked the elicitation coverage medians. Despite markedly fewer questions in Emergency cases, the elicitation fraction remained high, indicating that questioning—not volunteering—drove most surfaced normalized features.

### 3.3 User burden

CareRoute demonstrated urgency-conditioned questioning, asking substantially fewer questions for high-acuity cases: Emergency (median 10; IQR 4–14) vs. Doctor Visit (median 19; IQR 18–20) and Self Care (median 19; IQR 17–20).

#### 3.3.1 Time estimates

Using measured 99th percentile app response times (5 seconds) and estimated user input time (10 seconds per question), median session duration was 4.8 minutes overall (IQR 4.0–5.0), with notably shorter sessions for emergency cases (2.5 minutes; IQR 1.0–3.5) compared to Doctor Visit and Self Care cases (∼4.8 minutes each).

### 3.4 Case example: Kidney stones

#### Vignette

“A 45-year-old white man presents to the emergency department with a 1-hour history of sudden onset of left-sided flank pain radiating down toward his groin. The patient is writhing in pain, which is unrelieved by position. He also complains of nausea and vomiting.”

#### Condensed Format

“45 y/o m, 1 hour severe left-sided flank pain radiating into groin, nausea, vomiting, pain unrelieved by position”

#### Normalized features (9 items)

-45 y/o m - 1 hour - severe - left-sided - flank pain - radiating into groin - nausea - vomiting - pain unrelieved by position

#### Interactive session

*Presenting complaint:* “flank pain” - *Questions asked:* 16 - *Features elicited:* 8/9 (89%) - *Triage issued:* Emergency Care (matches reference triage)

## 4 Discussion

In this interactive evaluation, CareRoute achieved high triage concordance (88.9%) while minimizing user burden in high-acuity cases. Most normalized features were surfaced through questioning rather than volunteering, demonstrating that adaptive-questioning contributes meaningfully to safe triage.

### 4.1 Comparison with existing evaluation methods

Prior symptom checker evaluations fall into four categories, none of which adequately measure history-taking quality:

- **Static-input:** Pre-extracted symptom lists entered once, scoring only triage/diagnosis^1,7^
- **Interactive-but-unquantified:** Symptom checkers ask questions but studies don’t measure elicitation effectiveness^8^
- **Patient-based studies:** Real-world use with clinical adjudication, but still not isolating elicitation quality^9^
- **Retrospective reviews:** Re-analysis of cases focusing on diagnostic accuracy, not questioning strategy^2,3,10^

Reviews note that vignette-only evaluations may inflate performance and fail to capture real interaction dynamics^2,11,12^.

#### 4.1.1 Our methodological innovation

Our protocol uniquely measures three key dimensions:

1. Triage concordance/safety
2. History-taking quality: elicitation coverage and elicitation fraction
3. User burden (questions asked and time required)

This approach directly evaluates history-taking quality while demonstrating that urgency-conditioned questioning (fewer questions for emergencies) can maintain safety while improving efficiency.

### 4.2 Limitations

This study has the following limitations: (1) the modest sample (45 vignettes) limits precision and may limit generalizability; (2) a single evaluator ensures consistency but does not reflect varied user communication styles; (3) the behavior of generative AI-based applications can vary across versions and runs, and we did not assess robustness.

## 5 Conclusions

Our interactive evaluation protocol addresses a critical gap in symptom checker assessment by measuring both triage accuracy and history-taking quality. Using this approach, CareRoute achieved 88.9% triage concordance on the Semigran-45 benchmark with perfect safety on emergency cases, while demonstrating strong history-taking quality (median elicitation coverage 67%; elicitation fraction 70%) via adaptive-questioning.

CareRoute’s urgency-conditioned approach—asking fewer questions for high-acuity cases— demonstrates that symptom checkers can balance thoroughness with efficiency. These findings support incorporating history-taking quality metrics into standard evaluation protocols for symptom checkers.

## Data Availability

All data produced are available online at

https://github.com/healtheja/carerouteai-data

## 6 Data and materials availability

Per-case evaluation data, session transcripts, and code for metrics computation are available at https://github.com/healtheja/carerouteai-data. Reviewers can reproduce the protocol using the free CareRoute app (https://careroute.ai/get), version 0.3.71 or later.

## 9 Ethics statements

Not applicable (vignette-based study; no human participants).

## 10 Disclaimer

CareRoute is not a substitute for professional medical advice, diagnosis, or treatment. Users should seek professional medical advice for concerning symptoms.

## Appendix

**Table A1.**
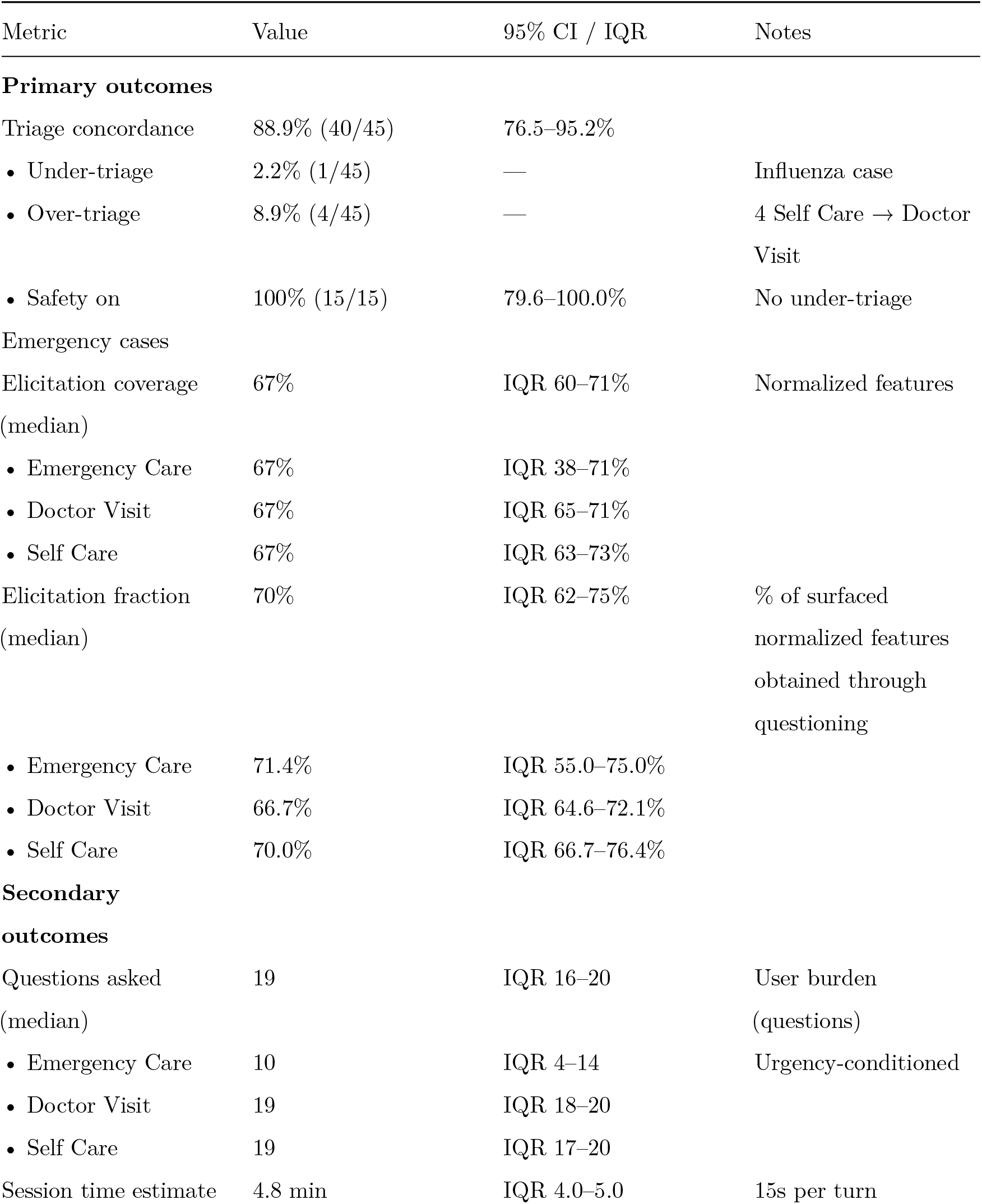
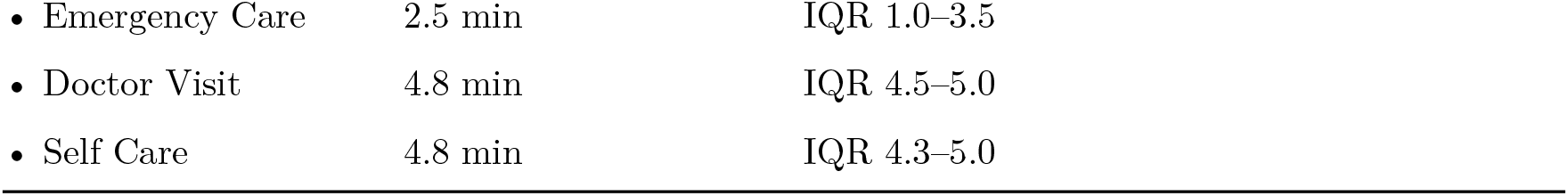
Summary of Evaluation Results.

